# The Development and Validation of the Global Post Trauma Symptom Scale-Uganda among Trauma-Affected Adults

**DOI:** 10.1101/2024.10.04.24314918

**Authors:** Lynn Murphy Michalopoulos, Melissa Meinhart, Erin Walton, David Robertson, Autumn Thompson, Thomas Northrup, Jong Sung Kim, Anne Conway, Nikita Aggarwal

**Author notes:** These authors contributed equally to this work (Lynn Murphy Michalopoulos, Melissa Meinhart, Erin Walton). These authors also contributed equally to this work (David Robertson, Autumn Thompson, Thomas Northrup, Jong Sung Kim, Anne Conway, Nikita Aggarwal).

## Abstract

The purpose of this study was to adapt and validate the Global Post-Traumatic Stress Scale (GPTSS) among adult caregivers of youth living with HIV in Uganda. This is the first study to adapt and validate a non-western instrument measuring post-traumatic symptoms in Uganda, which is critical in the accurate assessment of caregiver trauma on psychosocial functioning. The study utilized qualitative (i.e., cognitive interviews) and quantitative (e.g., classical test and item response theory) methods to establish content, criterion, and construct validity. The results indicated that the GPTSS was a valid and reliable assessment tool for the population. Specifically, cognitive interview results (n=31) indicated that participants broadly understood items of the GPTSS, but revisions in wording were needed to capture the intended meaning of some items. A reduced version of the GPTSS, GPTSS-U, was created through item response theory analyses based on discrimination criteria and difficulty parameters. A positive relationship between higher GPTSS-U and the PHQ-9 scores indicated acceptable criterion validity with a moderate effect size (r = 0.547; *p* < 0.000). Results from the graded response model indicated high discrimination parameters (range *b*= -.88, 3.14). Test information function curve findings indicated that the GPTSS is most precise at moderate to severe levels of post-traumatic symptoms. Using nested linear regression models, we found incremental validity as the total scores on the GPTSS-U significantly predicted functional impairment (*p* < 0.05) beyond the impact of the Post Traumatic Stress Checklist-5. Overall findings indicate that post-traumatic symptom measures must be adapted and developed for cross-cultural use to maintain validity and contextual relevance.

## Introduction

Exposure to potentially traumatic events can compound ongoing daily stressors, such as poverty and living with HIV. Among populations in low and middle-income countries, this is especially alarming as the risk of experiencing a traumatic event and developing a mental health disorder is higher due to contextual risk factors, such as poverty, conflict, and displacement (1–8). We are at a critical junction in global mental health research, as projections indicate the population of LMIC will double by 2053 (9), and awareness around trauma and global mental health continues to increase (8,10–13).

Unfortunately, most global assessments of post-trauma symptoms rely on Western assessment tools with a heavy emphasis on Post-Traumatic Stress Disorder (PTSD) (8,10,14–16). For example, Fodor et al.(8) found that 87% of research studies examining traumatic stress were in high-income countries (HIC), with over half of the papers (51%) being from researchers in the United States. As a latent construct, PTSD is similar among Western trauma-affected populations; however, there are inconsistent findings related to the relevance and reliability of PTSD among trauma-affected populations from LMIC (17,18). There is an ongoing debate in global mental health around PTSD, whereby some researchers assess PTSD as a standardized global disorder (7,19), and others emphasize local expressions of trauma (20–22).

In response to the lack of instruments that combine both perspectives, the Global Post-Trauma Symptom-Item Bank (GPTS-IB) was developed (7,23). The GPTS-IB is a 51-item bank based on a systematic review of qualitative studies that indicated post-trauma symptoms (24). The GPTS-IB acknowledges the potential utility of PTSD constructs in standard measures and accounts for other post-traumatic symptoms. The goal in creating the GPTS-IB was to establish an item bank that can be adapted and locally validated in various non-western trauma-affected settings. The application of GPTS-IB aims to make interpretations from norm-referenced scores about the perceived existence of symptomology related to ongoing trauma at the level of the individual.

Emerging research demonstrates that caregivers of children with high-need illnesses (e.g., HIV) often report traumatization from the caregiving experience. Caregivers reportedly have a higher prevalence of mental health needs (25,26), which can result in stigma and discrimination that can affect children under their care (27). Particularly relevant to the study context, caregivers in Uganda are often additionally burdened with insecure provisions for food and difficulties accessing health care (28).

For this study, the GPTS-IB was used to assess the nuances of post-traumatic symptoms among adult caregivers of children living with HIV in Uganda. The GPTS-IB scores were utilized to make inferences about the severity of post-traumatic symptoms experienced indirectly and directly among Ugandan caregivers of children living with HIV. The primary aim was to assess the use of mixed methods to reduce the GPTS-IB to a relevant scale among Ugandan adults (i.e., the Global Post Trauma Symptom Scale-Uganda; GPTSS-U). The secondary aim was to contextually validate the GPTSS-U using Classical Test Theory and Item-Response Theory (IRT). This is the first study aimed at utilizing the GPTS-IB to adapt and develop a reliable and valid post-trauma assessment tool in Uganda.

This study is especially significant for this region as most programs and services that address mental health and psychosocial functioning are now applied through task-shifting, a process where lay workers are trained to treat specific health disorders (29–31). As task-shifting approaches become more widely utilized to address the “treatment gap” in global mental health, research into its efficacy both for recipients of lay worker interventions and for their caretakers is needed (32); this will build upon the growing body of research to show the efficacy of treatment delivery by non-specialists (33–35). To our knowledge, this is the first study to validate a post-traumatic symptom instrument among trauma-affected caregivers of children with HIV in Uganda. Using the GPTSS-U in the caregiver context may demonstrate how mental health symptomology varies across cultural contexts, informing tailored responses to intervention development.

## Methods

### Background and Setting of Study

This study was conducted by the International Center for Child Health and Development (ICHAD) as part of two more extensive projects, SuubiI+Adherence and Suubi Maka, both in Uganda’s Rakai and Masaka regions. Suubi+Adherence is a longitudinal randomized control trial [NIH: 1R01HD074949-01] focusing on economic strengthening to increase adherence to HIV treatment for adolescents living with HIV in Uganda. Suubi Maka is a cluster randomized control trial that focuses on the educational outcomes of orphaned children through economic empowerment [NIH: RMH081763A]. Participants were included in the study if they were primary caregivers of a youth participant in the Suubi+Adherence or Suubi Maka study. The current study aimed to implement valid and reliable scales to measure psychosocial outcomes among caregivers within the interventions. Caregivers of youth living with HIV or orphaned because of HIV-related deaths were chosen as a vulnerable population at risk for multiple traumatic experiences and ongoing daily problems, indicating the likelihood that we would include participants both with and without post-trauma symptoms.

### Training

Ten local research staff members were trained to administer the psychosocial measures with caregivers of participants involved in the Suubi+Adherence or Suubi Maka study. All research staff received extensive training on cognitive interviewing methods and administration of quantitative assessment of psychosocial problems for the validation study. The research staff received ongoing feedback and training from the validation study’s principal investigator (PI) through role plays and group discussions among the training team.

### Sample

Participants were eligible to participate if they were indicated as caregivers of one or more intervention participants and over the age of 18. Exclusion criteria consisted of active psychosis or the presence of a significant developmental delay.

### Recruitment

Participants were recruited from February 1, 2016 until December 18, 2016.

### Consent and Ethical Approval

The cognitive interviews and validation study were approved by an IRB within a US-based and Uganda-based university. All procedures performed in this study involving human participants were in accordance with the ethical standards of the institutional and/or national research committee and with the 1964 Helsinki Declaration and its later amendments or comparable ethical standards. Written informed consent was obtained from all participants. Participants received UGX 10,000 (local currency, equivalent to USD 3.30) for the cognitive interviews and UGX 15,000 (USD 5) for the validation study.

### Measures

#### Demographics

Basic demographics of age, gender, and HIV status were assessed by self-report.

#### Global Post Trauma Symptom-Item Bank (GPTS-IB)

The GPTS-IB (23,36) was used to assess post-trauma symptoms. The GPTS-IB is a 51-item self-report item bank used to assess symptoms of post-trauma symptoms in adults. The GPTS-IB and subsequent reduced Global Post Trauma Symptom Scale-Uganda (GPTSS-U) use Likert scales to determine the frequency of symptoms experienced in the past two weeks. Response categories are based on four ordered response categories (*0 = none of the time*, *1 = a little of the time, 2 = most of the time,* and *3 = almost all of the time*). For each item of the GPTS-IB and GPTSS-U, participants were asked to indicate their experience of each symptom not in relation to a particular traumatic event.

#### Functioning Scale

A local measure of functional impairment, which was previously developed and validated among Ugandan adults in the Rakai and Masaka regions, was used in the current study (37). The measure includes tasks that men (9 items) and women (9 items) do to care for themselves, their families, and their communities (e.g., participate in community development activities, farm, care for children, etc.). Participants were asked how much difficulty they had performing each item in the past four weeks. Response options ranged from 0 ‘*no difficulty’* to 4 ‘*often cannot do*.’ Functional impairment was used as a validity criterion to assess the impact of post-trauma symptoms on participants’ daily lives.

#### Patient Health Questionnaire-9 (PHQ-9)

The PHQ-9 (38) is a nine-item self-report measure of symptoms of depression experienced in the past two weeks, with response options ranging from 0 ‘ *not at all*’ to 3 ‘*nearly every day*.’ The PHQ-9 is a commonly used measure of depression that has been used in non-Western contexts (39–43), including sub-Saharan Africa (44–46). In the current study, the PHQ-9 was used to determine the criterion validity of the GPTSS-U among participants because previous research has demonstrated support for the co-occurrence of depression and post-trauma symptoms among trauma-affected populations in both Western (47–50) and non-Western settings (51–53).

#### Post-Traumatic Stress-Disorder Checklist for DSM-5 (PCL-5)

The Post Traumatic Stress Disorder Checklist-5 (PCL-5; 54) is a 20-item questionnaire that corresponds with the DSM-5 symptom criteria for PTSD. In the current study, similar to the GPTS-IB, we asked participants to indicate how much each symptom bothered them in the past two weeks and used response categories *0 = none of the time*, *1= a little of the time, 2= most of the time* and *3 = almost all of the time,* rather than the five-category response options of the standard PCL-5. This was done to maintain continuity across assessment tools, which the study team believed would reduce confusion that could be caused by changing response categories. As such, the range for the PCL-5 total symptom severity is 0-60, which lowered the preliminary suggested cutoff score from 33 (54) to 20 in our study for a provisional PTSD diagnosis. The PCL-5 has been used among trauma-affected populations from LMIC (18,19,33,55). While the Post Traumatic Diagnostic Scale (56) has been validated among adolescents and young adults in Northern Uganda, it is based on the DSM-IV symptoms. The PCL-5 has not yet been validated in Uganda.

#### The Life Events Checklist for the DSM-5 (LEC-5)

The LEC-5 (54) is a 16-item questionnaire that is used to screen for exposure to potentially traumatic events (i.e., events associated with increased risk of PTSD or psychosocial distress) in a person’s lifetime. The LEC-5 has been widely used among trauma-affected populations from non-Western LMICs (55,57). In the current study, participants were asked to indicate if they had experienced or witnessed each potentially traumatic event. A total sum score was calculated to indicate the total number of events experienced (range 0-16) and witnessed (range 0-16), in addition to a total score of witnessed and experienced events (range 0-32).

### Procedure

#### Translation

All measures were translated into Luganda (the local language in the region) using translation-back translation, followed by group translation to ensure equivalence in the meaning of terms, conceptual understanding, and the ability of an adult with limited formal education to understand. Two translators certified in Luganda instruction– a secondary school teacher and a research assistant–completed the initial translation-back translation. The research team reviewed the translated assessment instruments and back-translated assessment tools item by item.

#### Data Analysis

To assess the face and content validity of the GPTS-IB, cognitive interviews were administered to 30 adult caregivers. Cognitive interviewing is a research method that establishes face and content validity by systematically asking participants how they comprehend items on a questionnaire (58,59). We specifically utilized the *think-aloud* cognitive interviewing method, which entails asking participants to describe each questionnaire item in their own words with examples (58). For example, participants were asked to describe the following in their own words and give examples for ‘*Feeling afraid, including being afraid that a traumatic event would happen again.*’ The administration included reading the 51 items of the GPTS-IB to the participant. All questions and responses were in Luganda, and answers were recorded verbatim using paper and pencil.

Responses to the *think-aloud* methods were assessed in terms of consistency with the intended meaning for each item. One author of the current study and a graduate-level research assistant coded responses of each item based on consistency with the intended meaning. Where the intended meaning was inconsistent, items were revised or changed for the validation study after full review from the research team for final validation before conducting the study.

#### Development of the GPTSS-U

For the validation study, the administration included reading each item of the measures to the participants. All responses were entered into Qualtrics (Qualtrics, 2016) by local research staff in Luganda. All research staff were fluent in both Luganda and English.

All data analyses were conducted using STATA 14 (Stata Corp, 2013). The GPTS-IB was reduced to a shortened and contextually specific version (GPTSS-Uganda; GPTSS-U) based on results from the item response theory analysis. We first conducted an exploratory factor analysis (EFA) to meet the assumption of unidimensionality (60). The assumption of unidimensionality is met if the factor analysis results in a 1-factor solution or a dominant first factor, with at least 20% of the variance explained (60–62). The assumption of local independence was tested through discrimination parameters for each item in the IRT model. Local independence was determined by a slope < 4.00 (60).

Based on the EFA results, a unidimensional graded response IRT model (GRM) was conducted to assess the degree to which the measure reflects the underlying latent variable (63). This study used the GRM as this model allows for ordered response categories. IRT was used to assess the ability of each item to differentiate among individuals at different points along the latent continuum of post-traumatic stress (64,65).

For each item, difficulty (or item location, *b*), discrimination (*a*), and measurement information (θ) parameters were estimated. Item difficulty parameters (*b*) indicate the amount of post-trauma symptoms where the probability of endorsing an item with a particular response category is 0.50. Three difficulty parameters (*b_1_, b_2_, b_3_*) were estimated, corresponding with the four possible response categories of the GPTS-IB. The first difficulty parameter (*b_1_*) indicates the level of the underlying latent trait of post-trauma symptoms, where the probability of endorsing an item with a “0” instead of “1” or “2” is 0.50. The second difficulty parameter (*b_2_*) is for the response of < 1. The third difficulty parameter (*b_3_*) is for the response of < 2. Discrimination parameters indicate the ability of a particular item to distinguish between individuals with low and high amounts of the latent construct of post-trauma symptoms. Values of 0.01-0.34 are considered to be very low, 0.35-0.64 low, 0.65 -1.34 moderate, 1.35 -1.69 high, and 1.70 and above very high (66). Measurement information (or precision) was determined by the Test Information Curve (TIC). Measurement information represents the certainty with which the measure assesses the underlying latent trait (θ) and can vary as a function of the level of θ.

We selected items for the GPTSS-U based on difficulty and discrimination parameters, as well as measurement information. Using item location estimates, we selected items that spanned a range of post-trauma symptom severity. We also selected items based on high discrimination values or with estimates of 1.35 and above. After selection of items for the GPTSS-U, we compared the TIC of the GPTS-IB to the GPTSS-U. We examined whether the selected items of the GPTSS-U had a similar TIC compared to the GPTS-IB across different levels of severity. Our final step included conducting a GRM IRT model to estimate item difficulty parameters and discrimination parameters of the reduced GPTSS-U scale.

### Data Analysis Plan for the Validation of the GPTSS-Uganda

#### Internal Consistency Reliability

Cronbach’s alpha scores were utilized to determine the internal consistency reliability of the GPTSS-U. A value between 0.70 and 0.79 is considered to be fair, 0.80 and 0.89 is considered good, and 0.90 and above is deemed excellent (67,68).

#### Construct Validity

Construct validity is the degree to which a scale measures the theoretical construct it was intended to measure and is associated with other related constructs. As such, construct validity was determined through Spearman’s correlation coefficients (*ρ*) to examine the strength of the relationship between the GPTSS-U and PHQ-9, PCL-5, trauma experienced from the LEC-5, and functioning scale. The effect size was based on a coefficient between 0.10-0.29, representing a small association, a coefficient between 0.30-0.49, representing a medium association, and a coefficient of 0.50 and above, representing a large association (69). We hypothesized that the GPTSS-U would be positively associated with each measure.

#### Criterion Validity

Criterion validity is defined as the relationship of a scale to a criterion variable, such as a clinical diagnosis (70). We used the Bolton method (37), a method used in LMICs to establish criterion validity where there are no psychiatrists available to provide psychiatric diagnostic assessments, to establish the criterion validity of the GPTSS-U. In the current study, we developed cases based on a PCL-5 cutoff score of 20 or above.

Non-cases were participants who had a score below 20 on the PCL-5. We then conducted a two-sample t-test with the hypothesis that cases would have a significantly higher score on the GPTSS-U than non-cases.

#### Sensitivity and Specificity Analyses

We conducted sensitivity and specificity analyses using receiver operating characteristic (ROC) curves to test the performance of the GPTSS-U and suggest appropriate cut-off scores for case identification. Similar to criterion validity, we used the cutoff of the PCL-5 as cases and non-cases. ROC curves plot the true positive rate (sensitivity) against the false positive rate (specificity). An area under the curve (AUC) of 0.50 indicates that the GPTSS-U would have no diagnostic utility, and an AUC of 1.0 would indicate perfect prediction. AUC values of 0.50–0.70 indicate low accuracy, 0.70–0.90 moderate accuracy, and above 0.90 high accuracy (71). An optimal cut-off point was generated for the GPTSS-U based on maximizing sensitivity and specificity (72).

#### Incremental Validity

Finally, we assessed the incremental or predictive validity of the GPTSS-U on functional impairment using nested linear regression models. In Model 1, we examined the impact of gender, age, HIV status, total LEC-5 traumatic events experienced, total PHQ-9 score, and total PCL-5 score. In Model 2, we added the GPTSS-U total score to the model. Incremental validity would be supported if total scores on the GPTSS-U significantly predicted functional impairment (*p* < 0.05) beyond the impact of the PCL-5. This was measured by a statistically significant increase in the R^2^ statistic when comparing Model 1 to Model 2 (73). To measure collinearity between scores on the PCL-5 and the GPTSS-U, we also examined the variance inflation factor (VIF). A VIF of 5 or above would indicate concern that the variables were highly collinear (74).

## Results

### Cognitive Interviews

Thirty participants (n=30) completed the cognitive interviews (83.33% female). The average age was 49.07 years (SD=11.18, range 33-83 years). Verbal responses from the cognitive interviews were back-translated into English, compiled in Excel, and coded by main themes. Responses from the think-aloud methods were coded and reviewed by two members of the research team in terms of consistency with the intended meaning for each item. The majority of items required slight revision in wording or additional description in meaning. For example, many participants described ‘*feeling detached from others’* as ‘*feelings of sorrow.’* As such, we revised the item with the addition of more details in the description, i.e., ‘*feeling detached or cut off from others’ (i.e., alienated, isolated, lonely, withdrawn or unable to socialize with others)*.

### Demographics for Validation Study

Two hundred caregivers (n=200) completed measures for the validation study (80.50% female). The average age of participants was 47.70 years (SD= 12.52, range 19-83).

### Creating the GPTSS-U from the GPTS-IB

#### Unidimensionality Assumption

A simple factor solution was obtained using Varimax rotation. Kaiser Meyer Olkin (KMO) measure of sampling adequacy was superb with .84 (75) and a significant Bartlett’s test of sphericity (χ2 (_1275_) = 4264.56, p<.001) suggesting a factor analysis was appropriate to conduct with the data. Seven factors were suggested from initial eigenvalues over 1. However, the first factor indicated over 44% of the variance, and, as such, unidimensionality was determined.

#### Local Independence

The graded response IRT model indicated local independence of the GPTS-IB as all discrimination parameters were < 4.00.

#### Selection of Items

The graded response IRT model indicated items with discrimination parameters ranging from low to high. For example, items such as ‘*headaches or migraines’* (*a* = 0.48), ‘*problems related to a long-term condition or a physical health problem*’ (*a* = 0.66), and ‘*unable to eat or lack of appetite*’ (*a* = 0.69) were indicated as poor in the ability to distinguish between participants with low and high amounts of post-trauma symptoms. Conversely, items such as ‘*sadness, sorrow or depressed mood’* (*a* = 1.86), ‘*emotional pain, suffering, despair or distress*’ (*a* = 1.97), and ‘*feeling confused or unable to figure out what was going on*’ (*a* = 2.10) were indicated as high in their ability to distinguish between participants with high and low amounts of post-trauma symptoms. Discrimination parameters for the GPTS-IB are presented in Table 1. Based on these estimates and item locations across low, moderate, and severe levels of post-trauma symptoms, the GPTS-IB was reduced to 11 items for the GPTSS-U.

**Table 1.** GPTS-IB Discrimination Parameters.

***GPTS-IB Discrimination Parameters***
| GPTS-IB Item | Discrimination Parameter<br>($a$) (Standard Error) |
| --- | --- |
| 1. Felt afraid, including being afraid that a traumatic event or stressful event would happen again. | 0.89 (0.16) |
| 2. Felt detached or cut off from others (i.e., alienated, isolated, lonely, withdrawn, or unable to socialize with others). | 0.93 (0.18) |
| 3. Had disturbed sleep or trouble sleeping (difficulty falling or staying asleep). | 0.85 (0.16) |
| 4. Been unable to experience positive emotions (i.e., happiness, joy, excitement). | 1.34 (0.20) |
| 5. Felt apathetic or that life has become meaningless | 1.19 (0.24) |
| 6. Had persistent negative expectations or felt hopeless. | 0.82 (0.18) |
| 7. Experienced repeated and disturbing memories about a traumatic or stressful event. | 1.64 (0.21) |
| 8. Felt agitated, irritated, frustrated, or angry. | 1.03 (0.18) |
| 9. Cried frequently. | 1.30 (0.24) |
| 10. Felt sadness, sorrow, or a depressed mood. | 1.9 (0.25) |
| 11. Experienced disturbing dreams or nightmares related to a stressful or traumatic event. | 1.08 (0.19) |
| 12. Experienced excessive worry, anxiety, or an ongoing sense of panic. | 1.35 (0.21) |
| 13. Had frequent arguments or fights with an intimate partner, family, or community member. | 0.90 (0.22) |
| 14. Had headaches/migraines. | 0.48 (0.15) |
| 15. Had trouble concentrating. | 1.13 (0.20) |
| 16. Felt forgetful or experienced memory loss. | 0.87 (0.17) |
| 17. Felt you were thinking too much. | 1.55 (0.21) |
| 18. Felt that no one understands what you have been through or are currently experiencing. | 0.89 (0.17) |
| 19. Had a lack of or reduced interest in activities that you used to enjoy. | 0.89 (0.16) |
| 20. Felt tired, fatigued, weak, or exhausted. | 1.04 (0.17) |
| 21. Experienced irritable behavior or angry outbursts. | 1.11 (0.19) |
| 22. Engaged in reckless, self-destructive, risky, or harmful behavior (i.e., abusing alcohol/drugs, risky sex, cutting). | 0.73 (0.33) |
| 23. Become emotionally upset when reminded of a stressful or traumatic event. | 1.48 (0.22) |
| 24. Felt a sense of loss. | 1.11 (0.19) |
| 25. Felt an ongoing sense of uncertainty, helplessness, or a lack of control. | 1.11 (0.18) |
| 26. Had thoughts of wanting to kill yourself. | 1.11 (0.18) |
| 27. Been unable to eat or a lack of appetite. | 0.76 (0.40) |
| 28. Been unable to talk about the trauma or feeling a sense of silence. | 0.69 (0.17) |
| 29. Felt that a traumatic or stressful event is actually happening again; that you are reliving it in the present (flashbacks). | 1.57 (0.23) |
| 30. Felt guilty. | 1.54 (0.23) |
| 31. Felt ashamed or damaged. | 1.28 (0.21) |
| 32. Felt jumpy or easily startled such as jumping or flinching in response to an unexpected noise. | 1.45 (0.24) |
| 33. Actively avoided external reminders of a stressful or traumatic event. | 1.06 (0.18) |
| 34. Experienced physical heart problems: heart palpitations, cardiovascular problems, or symptoms related to a heart condition. | 1.26 (0.21) |
| 35. Felt as if your surroundings or environment are not real. | 1.07 (0.18) |
| 36. Felt emotional pain (suffering), despair, or distress. | 1.14 (0.23) |
| 37. Experienced dizziness, felt confused, or disoriented. | 1.97 (0.26) |
| 38. Had an upset stomach or digestive problems. | 2.10 (0.28) |
| 39. Actively avoided thoughts, feelings, or memories related to a stressful or traumatic event. | 0.72 (0.16) |
| 40. Experienced a loss of faith and trust in the world, others, or spiritual beliefs. | 1.01 (0.18) |
| 41. Felt watchful of your surroundings or on guard. | 1.20 (0.22) |
| 42. Experienced physical trembling or shaking. | 1.08 (0.19) |
| 43. Felt chest pain or pressure in the chest. | 1.12 (0.21) |
| 44. Felt as if you have gone mad/crazy (i.e., seeing things that are not there). | 0.82 (0.17) |
| 45. Experiencing problems related to the heart (i.e., heartache, a weak heart, spoiled heart, dead heart, sick heart, tired heart, wounded heart, broken heart, heart in | 1.11 (0.32) |
| trouble, unclear heart, disturbed heart, falling heart, hot heart.) |  |
| 46. Experienced problems related to a long term condition or physical health problem (i.e., immune system disorders, diabetes, arthritis). | 1.16 (0.20) |
| 47. Had vision problems, eye problems, or weak/tired eyes. | 0.66 (0.17) |
| 48. Blamed yourself or others for a stressful or traumatic event. | 0.84 (0.16) |
| 49. Had an inability to remember an aspect or parts of a stressful or traumatic event. | 1.15 (0.19) |
| 50. Had a physical reaction when you were reminded of a stressful or traumatic event (i.e., sweating, racing heartbeat, nausea). | 1.40 (0.21) |
| 51. Having persistent and ongoing negative feelings such as anger, fear, guilt, horror, and sadness | 1.32 (0.21) |

### Descriptive Statistics for the GPTSS-U

Table 2 presents results from the GRM IRT model for the 11 selected items of the GPTSS-U. Discrimination parameters remained in the acceptable range for all items (*a =* 1.34 to *a* = 2.24), with item locations ranging from *b_1_* = -0.88 to *b_3_* = 3.14 across the latent construct. Test Information Curves for the GPTSS-U revealed little change from the GPTS-IB with both indicating the most amount of information at average to moderate levels of post-trauma symptoms (θ = 0 to θ = 2). Internal consistency reliability was good for the GPTSS-U with α = 0.87 and no improvement upon removal of any item. The average total score of the GPTSS-U among participants was 8.09 (SD = 5.92), with scores ranging from 0-29 among participants (scores could range between 0-33).

**Table 2.** Graded Response Model Item Discrimination Parameters (a) and Standard Errors, and Item Difficulty Parameters (b_1_, b_2_, b_3_,) and standard errors, N=200.

| GPTSS-U Item | Discrimination Parameter ($a$) (SE) | Difficulty Parameters ($b$) (SE) |
| --- | --- | --- |
| Experienced repeated, disturbing or unwanted thoughts or memories about a traumatic or stressful event | 1.82 (.25) | $b_1 = -.88 (.15)$<br>$b_2 = .66 (.13)$<br>$b_3 = 1.97 (.24)$ |
| Felt sadness, sorrow or a depressed mood | 2.19 (.31) | $b_1 = -.58 (.12)$<br>$b_2 = 1.14 (.15)$<br>$b_3 = 2.18 (.25)$ |
| Experienced excessive worry, anxiety or an ongoing sense of panic | 1.49 (.23) | $b_1 = -.07 (.13)$<br>$b_2 = 1.41 (.21)$<br>$b_3 = 2.56 (.37)$ |
| Feeling you were thinking too much | 1.62 (.24) | $b_1 = -.61 (.14)$<br>$b_2 = 1.18 (.18)$<br>$b_3 = 2.17 (.29)$ |
| Becoming emotionally upset when reminded of a stressful or traumatic event | 1.51 (.24) | $b_1 = .02 (.13)$<br>$b_2 = 2.01 (.29)$<br>$b_3 = 2.97 (.44)$ |
| Been unable to talk about a traumatic or stressful event or feeling a sense of silence | 1.49 (.24) | $b_1 = .06 (.13)$<br>$b_2 = 2.14 (.30)$<br>$b_3 = 2.52 (.36)$ |
| Feeling that a stressful or traumatic event is happening again; that you are reliving it in the present (flashbacks) | 1.60 (.26) | $b_1 = .26 (.13)$<br>$b_2 = 1.66 (.23)$<br>$b_3 = 2.44 (.34)$ |
| Feeling ashamed or damaged | 1.45 (.25) | $b_1 = .65 (.15)$<br>$b_2 = 2.05 (.31)$<br>$b_3 = 3.14 (.52)$ |
| Feeling emotional pain<br>(suffering), despair or distress | 2.02 (.30) | $b_1 = -.29 (.12)$<br>$b_2 = 1.34 (.17)$<br>$b_3 = 2.12 (.26)$ |
| Feeling confused or unable to<br>figure out what was going on | 2.24 (.33) | $b_1 = -.05 (.11)$<br>$b_2 = 1.35 (.16)$<br>$b_3 = 2.16 (.26)$ |
| Had an inability to remember<br>parts of a stressful or traumatic<br>event | 1.34 (.22) | $b_1 = .10 (.14)$<br>$b_2 = 2.13 (.32)$<br>$b_3 = 2.83 (.43)$ |

#### Construct Validity of the GPTSS-U

A positive relationship between higher GPTSS-U and the PHQ-9 scores indicated construct validity with a moderate effect size (*r* = 0.54 *p* < 0.001) and a strong effect size between the GPTSS-U and the PCL-5 (*r* = 0.91 *p* < 0.001). In addition, although effect sizes were very small, GPTSS-U scores and trauma experienced (*r* = 0.17), trauma witnessed (*r* = 0.04), and trauma experienced or witnessed (*r* = 0.14) were also significantly positively associated (*p* < .05). GPTSS-U scores and function scale scores were also correlated with a moderate effect size (*r =* .304 *p* < 0.001). Table 3 displays the polychoric correlation matrix for (1) GPTSS-U, (2) PHQ-9, (3) trauma experienced, (4) trauma witnessed, (5) trauma total, and (6) functional impairment scale. A very strong correlation between the GPTSS-U and PCL-5 (r=0.9, *p*<0.001) and a strong correlation between the GPTSS-U and PHQ-9 (r=0.54, *p*<0.001) support the construct validity of the GPTSS-U.

**Table 3.** Polychoric Correlation Matrix.

| Variable | GPTSS-U | PHQ-9 | Trauma experienced | Trauma witnessed | Trauma Total | PCL-5 | Functioning Scale |
| --- | --- | --- | --- | --- | --- | --- | --- |
| GPTSS-U | 1.00 |  |  |  |  |  |  |
| PHQ-9 | 0.54*** | 1.00 |  |  |  |  |  |
| Trauma Experienced | 0.19** | 0.19** | 1.00 |  |  |  |  |
| Trauma Witnessed | 0.03* | -0.04 | -0.04 | 1.00 |  |  |  |
| Trauma Total | 0.15* | 0.09 | 0.62*** | 0.75*** | 1.00 |  |  |
| PCL-5 | 0.90*** | 0.58*** | 0.15* | -0.04 | 0.06 | 1.00 |  |
| Functioning Scale | 0.31*** | 0.20** | 0.61*** | 0.70*** | 0.95*** | 0.26*** | 1.00 |

*Note:* GPTSS-U = Global Post Trauma Symptom Scale - Uganda; PHQ-P = Patient
Health Questionnaire-9 ; PCL-5 = Post-Traumatic Stress-Disorder Checklist for DSM-5.
\* $p<0.05$ ; \*\* $p<0.01$ ; \*\*\* $p<0.001$

#### Criterion Validity of the GPTSS-U

Results from the two-sample t-test with unequal variances established criterion validity of the GPTSS-U scale with cases (n=45) having a significantly higher mean (15.96, SD = 4.74) than the mean (5.8, SD = 3.95) of non-cases (n=155) (*p* < 0.001).

#### Predictive Validity

The results of the incremental validity analysis are presented in Table 4. Nested regression was used to compare the Models 1 and 2. Both models analyzed the effects of (1) gender, (2) age, (3) HIV status, (4) traumatic experiences, (5) symptoms of depression, and (6) symptoms of PTSD on functional impairment. Model 2 included mean GPTSS-U scores in the analysis. Standardized beta coefficients were reported for all models. Forty-three percent of the total variance in impaired functioning was explained by Model 2,, which represented a 0.13% change in R^2^ (explained variance) with the addition of the average GPTSS-U scores to the final model. The results of Model 2 indicated that *ceteris paribus*, a one-unit increase in average GPTSS-U was associated with a 0.266 increase in impaired functioning. The VIF for GPTSS-U was 1.72, and the tolerance was 0.58 showing no multicollinearity. Notably, when GPTSS-U scores were included in Model 2, the association between PCL-5 and impaired functioning became non-significant. An *F* test of the change in R^2^ when GPTSS-U was added to Model 2 was statistically significant (*p* <0.05), indicating further support for the predictive validity of the GPTSS-U. Models 3 and 4 serve as sensitivity analyses by reverse nesting the model to first include the GPTSS-U score in Model 3 and adding in the PCL-5 score as a comparator in Model 4. This approach did not demonstrate a change in explained variance, and the association between GPTSS-U and impaired functioning maintained its statistically significant association.

**Table 4.** Effects of Measured Variables on Impaired Functioning Presented as Beta Coefficients.

| Model | $\beta$ (S.E) | *t* |
| --- | --- | --- |
| Model 1 |  |  |
| Gender | 0.047 (0.89) | 0.85 |
| Age | 0.114 (0.03) | 1.95 |
| HIV | 0.04 (0.73) | 0.7 |
| Trauma | 0.59 (0.18) | 10.46** |
| PHQ-9 | 0.01 (0.11) | 0.13 |
| PCL-5 | 0.148 (0.05) | 2.15* |
| Model 2 |  |  |
| Gender | 0.034 (0.89) | 0.85 |
| Age | 0.116 (0.03) | 2.01* |
| HIV | 0.042 (0.72) | 0.73 |
| Trauma | 0.576 (0.18) | 10.24** |
| PHQ-9 | 0.002 (0.11) | 0.03 |
| PCL-5 | -0.086 (0.1) | -0.66 |
| GPTSS-U | 0.266 (0.14) | 2.09* |
| Model 3 |  |  |
| Gender | 0.039 (0.88) | 0.7 |
| Age | 0.112 (0.03) | 1.95 |
| HIV | 0.041 (0.72) | 0.73 |
| Trauma | 0.578 (0.17) | 10.33** |
| PHQ-9 | -0.009 (0.11) | -0.14 |
| GPTSS-U | 0.195 (0.07) | 2.94* |
| Model 4 |  |  |
| Gender | 0.034 (0.89) | 0.61* |
| Age | 0.116 (0.03) | 2.01 |
| HIV | 0.042 (0.72) | 0.73 |
| Trauma | 0.576 (0.18) | 10.24** |
| PHQ-9 | 0.002 (0.11) | 0.03 |
| GPTSS-U | 0.266 (0.12) | 2.09* |
| PCL-5 | -0.086 ( 0.1) | -0.66 |

*Note:* GPTSS-U = Global Post Trauma Symptom Scale - Uganda; PHQ-P = Patient
Health Questionnaire-9 ; PCL-5 = Post-Traumatic Stress-Disorder Checklist for DSM-5
(PCL-5)
\* $p < 0.05$ ; \*\* $p < 0.001$

#### Clinical Utility

The GPTSS-U had an area under the curve (AUC) of 0.95 (95% CI 0.92-0.98) when post-traumatic stress symptoms were compared to an absence of symptoms. The cut-off score that was determined to be ideal for the GPTSS-U was 11, which aligned with a sensitivity of 88.9% and specificity of 87.7% for post-traumatic stress symptoms compared with an absence of symptoms.

## Discussion

This study aimed to adapt and test the psychometric properties of the GPTSS and GPTSS-U among caregivers of youth living with HIV in Uganda, utilizing both qualitative and quantitative methods. The results demonstrate the effectiveness of mixed methods in adapting and validating the scale, demonstrating the relevance of the adapted GPTSS-U within the Ugandan context. Discrimination parameters were all in the high range (see Table 2), indicating the scale’s ability to distinguish between high and low levels of post-traumatic stress. The graded response model also demonstrated various difficulty parameters across questions. The difficulty parameters spanned the latent construct, which means that the items in the scale represent expressions of the latent construct with low, moderate, and high levels of difficulty to identify. The study also found that the GPTSS-U is a reliable measure with demonstrable content, criterion, and construct validity for caregivers of youth living with HIV in Uganda.

While not intended for diagnostic accuracy, the GPTSS-U provides additional evidence for the nuances of mental health symptomatology across contexts. Specifically, our results indicated that the GPTSS-U is a better predictor of functioning among Ugandan caregivers compared to the standard PCL-5. Further, using IRT allowed us to determine how precise the scale is at different levels. These findings showed that the GPTSS-U was most accurate at identifying moderate and severe levels of post-traumatic stress. Therefore, the GPTSS-U could help identify those with post-trauma symptoms who are most in need of interventions. Further, the test information function curve confirms that the GPTSS-U gives the most information or precise measurement at moderate to severe levels of post-traumatic stress.

The average total score of the GPTSS-U across all participants was 8.09 (SD = 5.92), indicating that most participants reported mild to moderate severity of post-trauma symptoms. A positive relationship between higher GPTSS-U and PHQ-9 scores indicated acceptable criterion validity and provided additional evidence for the utility of the GPTSS-U. Additionally, items from the item bank with the lowest discrimination were related to physical health, which is contextually sound and may be related to the difficulty in disentangling the cause of physical health problems from stress and mental health.

Item response theory (IRT) was used in conjunction with Classical Test Theory (CTT) to explore the relationship between the latent trait (trauma) and the nuances of its symptomatology. Using cognitive interviews as a first step in the validation process demonstrated high levels of utility in establishing semantic equivalence. Although post-traumatic symptoms have been noted to be a relevant concept among caregivers globally (25,27) and in Uganda (28), the use of cognitive interviews was critical in understanding how post-traumatic symptoms are defined and understood in context. These findings support the need to assess content validity before administering the GPTSS in different contexts. Content validity should also be conducted before future application of the GPTSS-U, especially among different populations within Uganda.

Findings supported using quantitative and qualitative methods to establish validity and ensure culturally relevant contextual application. Examples of validating mental health instruments exist among other sub-Saharan African populations. Michalopoulos et al. (36) adapted and validated a measure of HIV-related shame, the Shame Questionnaire (SQ), among Ugandan youth living with HIV using a similar methodology. The SQ was also tested for validity and reliability among traumatized girls in Lusaka, Zambia (18) using classical test and item response theory methods. Applying the SQ across populations demonstrated its general utility and the need for additional, similar studies in LMICs. The International Depression Symptom Scale-General (IDSS-G) is used to screen for mental health disorders in LMICs while eliminating the Western bias embedded in most self-report measures (32). It was found to be a valid and reliable measure, with the ability to accurately predict functional impairment in this population.

Although we utilized methodology that is similar to previous cross-cultural validation studies, our study is the first known measure that was developed and validated to examine post-trauma symptoms among Ugandan caregivers. Although the GPTSS was also deemed valid and reliable among other populations globally (23), comparing the results indicates a difference in item difficulty parameters across the latent post-traumatic symptoms construct. Such differences could be due to varying cultural contexts or changes in the wording of some of the items of the current study. Regardless, the differences in item location highlight the importance of future validation studies in non-western contexts, which should include examination of differences by gender, age, location, and type of trauma experienced.

An additional limitation of this study is that the sample represents the greater Masaka region. Future studies should examine the utility of the GPTSS-U and differential item functioning across varying regional contexts in Uganda and other countries in Sub-Saharan Africa with a high prevalence of HIV. Through further study, researchers can consult with local health and mental health professionals, caregivers, and additional family members to gauge holistically whether adjustments to the language GPTSS-U may be appropriate given cultural attitudes and understandings of symptomatology.

According to research in this area, the sample size was adequate (32,60). However, our sample size did not allow for the disaggregation between demographic characteristics. Of primary interest would be investigating gender differences in post-traumatic symptoms. Research has shown inconsistent results comparing trauma symptoms (specifically PTSD) between males and females, while other research shows invariance in the factor structure of PTSD (76–78). Even after controlling for PTSD severity, other studies found differences between genders in item endorsement (79,80). For example, women were more likely to report feeling more emotionally distant than men (79). Future research needs to examine post-traumatic symptoms across genders, which was not feasible for this study due to the smaller sample size.

Although difficulty parameters spanned the whole construct, some items clustered in the middle overlapped. While the item information function indicated potential overlap of some items, we retained all items to ensure acceptable reliability. Future studies could explore reducing the overlapping categories where potential redundancy exists to increase precision. However, this strategy was not applied to this study because the findings point to the validity of the overall scale.

This study demonstrates several strengths, including providing nuanced information on the mental health of an underrepresented population within global research, supporting advocacy efforts around contextually appropriate and effective mental health assessment tools, and increasing conversation on caregiver trauma exposure in Sub-Saharan Africa. Additionally, the awareness and dialogue with locally impacted communities alluded to themes of grief and loss related to caregiving and chronic illness, which encourages further conversation and study of the GPTSS scale efficacy.

## Conclusions

The findings of this study suggest that caregiving for individuals living with HIV is correlated with posttraumatic outcomes in Uganda and may be relevant to other countries in Sub-Saharan Africa, especially in regions with high HIV prevalence. The utility of the GPTSS-U indicates the high importance of further examination of contextual factors that may affect the presentation and expression of post-traumatic symptoms, such as gender, socioeconomic status, social support, region, caregiving responsibilities, and additional types of trauma. Further, the mixed methods, in addition to the utilization of both CTT and IRT to develop the GPTSS-U may be helpful in the development of relevant scales in contexts where validated scales are unavailable.

The results of this study indicate a critical need both for the development and implementation of evidence-based interventions to support caregivers of people with high-needs health issues and to inform best practices regarding post-traumatic stress interventions. Relevant interventions should be mindful of factors influencing the presentation and perpetuation of post-traumatic symptoms for this population, with particular emphasis on those tailored for individuals whose social determinants of health (e.g., greater economic instability) are most closely linked to post-traumatic stress disorder (81).

## Data Availability

Data will be fully available if accepted

